# Evaluating Large Language Models for ADHD Education: A Comparative Study of ChatGPT-5, DeepSeek V3, and Grok 4

**DOI:** 10.1101/2025.10.12.25337841

**Authors:** Xingmin Han, Ruirui Xing, Mi Zhou

## Abstract

**Background:** Children with attention-deficit/hyperactivity disorder (ADHD) often face barriers to participating in organized sports, particularly when physical education (PE) is delivered by outsourced coaches with limited training in disability inclusion. Meanwhile, large language models (LLMs) such as ChatGPT, DeepSeek, and Grok are increasingly used to generate educational content, yet their readability, stability, and accuracy for non-specialist educators remain unclear.

**Methods:** This study systematically compared three advanced LLMs, ChatGPT-5, DeepSeek V3, and Grok 4, using identical prompts related to ADHD definitions, symptoms, and medication–exercise interactions. Thirty responses per model were collected and analyzed for content accuracy, readability (Flesch–Kincaid Reading Ease, Grade Level, and SMOG), and lexical complexity.

**Results:** All models aligned with DSM-5 in describing ADHD but differed in emphasis and stability. DeepSeek V3 produced the broadest and most variable outputs, Grok 4 showed the greatest consistency and clinical structure, and ChatGPT-5 generated concise and strengths-based explanations. However, all models exhibited high reading levels (FKGL > 12), exceeding recommended public-health standards.

**Conclusion:** While LLMs demonstrate strong potential for generating ADHD-related educational materials, their current readability and stability limitations restrict accessibility for non-specialist educators. Future work should focus on optimizing prompt design and language calibration to enhance usability in inclusive education contexts.

## Introduction

Attention-deficit/hyperactivity disorder (ADHD) is one of the most prevalent neurodevelopmental disorders among children worldwide (1). It is characterized by persistent patterns of inattention, hyperactivity, and impulsivity (2). These difficulties often extend into adolescence and adulthood, with long-term implications for academic achievement, social interaction, and daily functioning (3).

Research has increasingly highlighted the role of physical activity and exercise in mitigating ADHD symptoms. Moderate-intensity aerobic exercise has been shown to alleviate core symptoms of ADHD (4–7). However, children with ADHD frequently encounter obstacles when engaging in organized sports (8). Evidence from motor control studies indicates that individuals with ADHD struggle to process long-duration or sequential information, resulting in irregularities in movement performance (9). In addition, these children are often less readily accepted by peers, particularly in structured activities that require cooperation and sustained attention, which further exacerbates their exclusion from group-based exercise opportunities (8). Consequently, participation in organized after-school activities frequently requires substantial teacher support. However, physical education (PE) teachers themselves often report feeling underprepared to meet the needs of children with disabilities, including ADHD (10). The situation in Australia is particularly noteworthy: a large proportion of primary school PE is outsourced to external providers, reaching as high as 78.1% in Victoria (11). The competence and preparation of these outsourced coaches, especially regarding knowledge of ADHD have been questioned (12). This raises critical concerns about whether children with ADHD are able to fully derive the intended developmental and psychosocial benefits of participation in organized after-school sports. Urgent interventions are required to address this issue.

Advances in artificial intelligence (AI) and large language models (LLMs) are beginning to reshape education. Cutting-edge models such as ChatGPT are increasingly applied to support teachers (13), clinicians (14) and parents (15) in understanding developmental disorders (16). Recent research has begun to explore the potential of LLM-generated educational materials for professional development, with the aim of equipping educators and caregivers with actionable strategies to improve children’s well-being (17). Nevertheless, prior studies have primarily focused on educators with specialized training, such as early childhood teachers or special education practitioners, who already possess a theoretical foundation in developmental disorders (14, 18). Far less attention has been paid to how LLMs may support those with no prior knowledge, such as parents, after-school sports coaches or casual PE instructors. They are widely recognized as lacking proper teaching qualifications and demonstrating limited awareness of ADHD (12, 19).

For these groups, educational materials must be highly readable, concise, and practically oriented to guide the design and delivery of after-school activities. Moreover, given their limited background knowledge, it is critical that generated materials are stable, accurate, and free from misleading content, as these users may lack the expertise to verify correctness. Yet, to date, no study has systematically examined the readability, accuracy, and usability of LLM-generated ADHD-related content for this population of educators.

Moreover, while multiple LLMs are currently available, each possesses distinct architectures, training approaches, and knowledge integration capacities, which may influence the clarity, comprehensiveness, and accessibility of the educational material they produce. For instance, ChatGPT-5 emphasizes human-like conversational adaptability (20), DeepSeek V3 prioritizes structured and analytical outputs (21), and Grok 4 is designed for rapid information retrieval and humor-infused explanations (22). Identifying the most effective mode for generating ADHD-related educational materials is crucial for guiding the future application of AI tools in inclusive education.

The present study aims to systematically evaluate the outputs of three leading LLMs, ChatGPT-5, DeepSeek V3, and Grok 4, to determine how effectively they can provide clear, accessible, and practical ADHD related knowledge. This study serves as a preliminary investigation within a broader series of AI related research, laying the groundwork for subsequent studies. Future work will involve interviews, application development, and empirical validation in the real contexts.

## Methods

### Study design

This study employed three large language models ChatGPT-5 (20), DeepSeek V3 (21), and Grok 4 (22) to generate educational materials. The educational context of outsourced physical education coaching was selected as the testing domain. As ChatGPT-5 adapts and optimizes its responses based on the user’s IP address (OpenAI, 2025), we report that all responses in this study were generated in Melbourne, Australia. The study was conducted from 7 August to 8 August 2025.

### Prompts

The prompts were designed to elicit accessible, user-friendly medical information from the AI system, following the approach described by Akkan and Seyyar (23) and Zhou, Pan (24). The three guiding questions were shown in Table 1

**Table 1.**
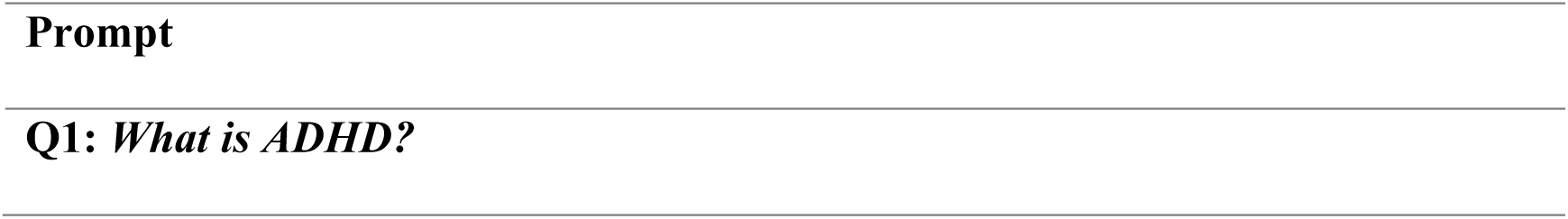
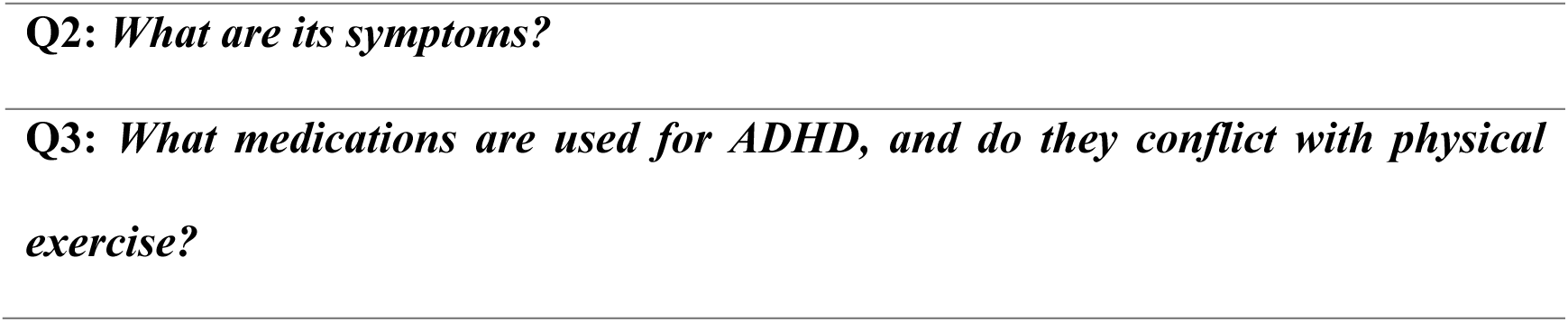
Questions for ADHD information.

### Procedure

Each prompt was administered to the three large language models under identical training and testing conditions (25). For each model, prompts were presented sequentially to ChatGPT 5, DeepSeek V3 and Grok 4. The browser cache was cleared and a new chat session was initiated, in line with best practices for minimizing memory and context carryover (26). All sessions were conducted on the same computer using a custom user account within a newly created virtual machine to ensure consistency of testing conditions (27). No additional questions or distracting content were introduced beyond the predefined prompts. The collected responses were copied into unformatted plain text files for subsequent analysis. To capture variability across runs, ten independent attempts were generated for each question, yielding a total of 30 responses per model (10 per question). Each question was entered independently, and the three models provided responses under identical conditions, without any additional clarifying prompts or contextual information.

## Measures

### Quality analysis

A qualitative content analysis was conducted by summarizing and comparing the presence of predefined key content elements within each response across four dimensions: language level, information depth, structural clarity, and additional content. Two reviewers (XMH and MZ) independently evaluated all responses, and any discrepancies were resolved through discussion with a third researcher (RRX).

### Readability

The readability of the responses was assessed using three well-established metrics: the Flesch–Kincaid Reading Ease (FKRE), the Flesch-Kincaid Grade Level (FKGL), and the Simple Measure of Gobbledygook (SMOG) (28, 29). The FKRE assigns a numerical score ranging from 0 to 100, with higher values indicating simpler and more accessible text. A score near 100 suggests that the content is very easy to understand, whereas lower scores denote increased complexity. The FKGL is an adaptation of the FKRE that estimates the minimum education level required for comprehension, expressed in U.S. school grade levels (28). A higher FKGL score corresponds to more complex text, implying that individuals with lower levels of formal education may find the material difficult to understand (28). The SMOG index also estimates the years of education needed for comprehension, but it focuses on the frequency of polysyllabic words (three or more syllables) within a standard 30-sentence sample (29). Higher SMOG scores indicate greater complexity and are particularly sensitive to technical or specialized vocabulary. Number and percentage of complex words in each response were also reported. All readability metrics were calculated using the WebFX online readability test (30).

### Data Analysis

A qualitative content analysis was conducted in a descriptive manner. The results for each model were summarized concisely in tabular form, while detailed analyses of individual responses are provided in the appendix. The quantitative analysis included the calculation of readability indices (Flesch Reading Ease, Flesch–Kincaid Grade Level, and SMOG), along with descriptive statistics for the number and proportion of complex words. All statistical analyses and visualizations were performed in R (version 4.3.2). Descriptive statistics and line plots were used to compare results across attempts, questions, and models.

## Results

### Content analysis

Table 2 illustrates the characteristics of the three models in their responses regarding the definition of ADHD (Q1). All three defined ADHD as a neurodevelopmental disorder characterized by inattention, impulsivity, and hyperactivity, and each referred to the three DSM-5 subtypes: Predominantly Inattentive, Predominantly Hyperactive-Impulsive, and Combined Type. This is consistent with DSM-5 (2). However, clear differences were observed among the three models. DeepSeek V3 highlighted clinical evaluation procedures (psychiatric/psychological assessments, symptom checklists, and multi-informant reports) and stressed the importance of ruling out other conditions.

**Table 2.** Comparative Analysis of the 10 Response for (Q1: What is ADHD?)

Grok-4 explicitly listed DSM-5 diagnostic requirements, including symptom persistence for ≥6 months, onset before age 12, and occurrence across multiple settings. GPT-5, by contrast, gave less attention to diagnostic criteria and instead emphasized genetic and neurotransmitter mechanisms, as well as differences in ADHD presentation between children and adults.

Moreover, both DeepSeek V3 and Grok-4 underscored a multimodal approach involving medication, behavioural therapy, and lifestyle adjustments. GPT-5 concluded by emphasizing that individuals with ADHD may also demonstrate creativity and vitality. Supplementary features further distinguished the models: DeepSeek V3 included a “Myths vs. Facts” section to address societal misconceptions, Grok-4 provided epidemiological data (5–10% of children, 2–5% of adults) and noted underdiagnosis among females, and GPT-5 highlighted the potential strengths of ADHD (creativity, energy, innovation) while suggesting a narrative approach to capture patients’ lived experiences.

Table 3 illustrates the characteristics of the three models in their responses regarding ADHD symptoms (Q2). All three models described ADHD symptoms in terms of inattention and hyperactivity/impulsivity, consistent with DSM-5 diagnostic criteria. Grok-4 explicitly cited DSM-5 as its reference point, and each model elaborated on the two core symptom domains.

**Table 3.** Comparative Analysis of the 10 Response for (Q2: What are the Symptoms of ADHD?)

DeepSeek V3 included a dedicated section on “ADHD in Adults,” highlighting chronic procrastination, disorganization, emotional instability, and impulsive spending as common features. Grok-4 emphasized developmental differences, noting that hyperactivity in children often manifests as running or climbing, whereas in adults it presents as “inner restlessness.” It also provided examples of comorbidities and daily life impacts, such as overlap with anxiety and depression, children forgetting homework, and adults making impulsive financial decisions. GPT-5 similarly stressed that adult hyperactivity is more likely to manifest as psychological unease rather than overt physical activity.

Table 4 illustrates the characteristics of the three models in their responses regarding whether ADHD medications interfere with physical exercise (Q3). All three models classified medications into stimulants and non-stimulants. They differed, however, in the level of classification detail, explanations of exercise-related risks, and the extent of supplementary information. DeepSeek V3 and Grok-4 provided the most comprehensive pharmacological breakdown, whereas GPT-5 presented a more concise classification. All models explained the mechanisms of action and emphasized that

**Table 4.** Comparative Analysis of the 10 Response for (Q3: What medications are used for ADHD, and do they conflict with physical exercise?)

ADHD medications generally do not conflict with physical exercise, though certain risks should be considered. Each recommended monitoring heart rate and blood pressure, along with maintaining adequate hydration.

Differences emerged in the scope of their recommendations. Grok-4 offered authoritative but relatively brief advice, acknowledging the benefits of exercise for ADHD management and suggesting a few activities. DeepSeek V3 provided the most detailed and pragmatic guidance, including specific exercise recommendations (e.g., jogging, swimming, yoga, strength training) and cautionary notes for high-risk activities (e.g., marathons, extreme HIIT, heat exposure). GPT-5, by contrast, did not include direct exercise-related recommendations, restricting its discussion to medication effects and general safety considerations.

### Readability and complexity

The model that exhibited the greatest variability was DeepSeek. Despite its average performance being in the mid-range, its fluctuations were pronounced in both readability and lexical complexity. In the Flesch–Kincaid Reading Ease, results across ten trials for Q1 ranged from 5.2 [95% CI: 3.8–6.7] to 27.6 [95% CI: 24.1–31.2], reflecting considerable instability, particularly in readability. In Q3, the score dropped sharply to –23.4 [95% CI: –26.9 to –19.8] before rising again to 5.1 [95% CI: 3.2–7.0]. The Number of Complex Words ranged from 115 to 229, while the Percentage of Complex Words remained between 30–38%.

GPT showed the second highest level of fluctuation, following DeepSeek. Its Flesch– Kincaid Reading Ease in Q1 ranged from 12.5 [95% CI: 11.0–14.1] to 19.8 [95% CI: 18.2–21.3]. Moreover, across the Flesch–Kincaid Grade Level, Gunning Fog Score, and SMOG Index, scores consistently increased in complexity, culminating in Q3 with a SMOG Index of 25.4 [95% CI: 23.7–27.0]. This indicates a steadily professional tone, though at the cost of reduced accessibility. In terms of lexical complexity, GPT was comparatively stable, with the Number of Complex Words ranging from 70–110 and the Percentage of Complex Words holding steady at 22–28%.

In contrast, Grok 4 was the most stable of the three models. Although its scores were consistently the lowest, with Flesch–Kincaid Reading Ease in Q2 ranging from 23.6 [95% CI: 21.8–25.5] to 32.2 [95% CI: 29.1–35.4], this suggests that its professional depth was somewhat lower than the other models. Nonetheless, its stability prevented the sharp readability swings observed in DeepSeek. Interestingly, Grok 4 presented the highest lexical complexity in Q3, with the Number of Complex Words peaking at approximately 230.

## Discussion

### Overview of research findings

All three models aligned with DSM-5 in defining ADHD and describing core symptoms but differed in scope, depth, and style. DeepSeek V3 was the most expansive, offering broad contextual details and practical guidance. Grok-4 was the most clinically structured and stable, closely following DSM-5 but with less breadth. GPT-5 was concise and accessible, highlighting strengths-based perspectives and emotional dimensions while showing moderate complexity. Readability analyses further showed that DeepSeek V3 had the greatest variability, GPT-5 displayed steadily increasing complexity, and Grok-4 remained the most stable and comparatively less complex.

### Comparison with previous studies

Berrezueta-Guzman, Kandil (14) showed that ChatGPT-5 can provide personalized guidance for children with ADHD, particularly by improving empathy, adaptability, and communication in simulated clinical environments. Similarly, Fuermaier and Niesten (31)assessed GPT-based responses in simulated ADHD scenarios, confirming their alignment with DSM-5 diagnostic standards and their ability to deliver structured definitions, symptom descriptions, and complementary recommendations. The findings of this study are consistent with these studies, emphasizing the increasing role of LLMs in inclusive education and support Our findings further extend the existing literature by evaluating multiple LLMs under identical conditions to examine the stability of response generation. This metric is particularly important when considering large-scale deployment, especially in contexts where end-users may lack the capacity to discern factual inaccuracies. Our results indicate that Grok-4 demonstrated the highest stability, suggesting its potential utility for educational settings. However, like GPT-5, it is subject to access restrictions. Frequent use within a single day requires a paid subscription, which may increase implementation costs. By contrast, DeepSeek V3 remains freely accessible.

Despite these advancements, the results also expose significant challenges. Consistent with earlier studies, we found that readability remains a key obstacle to the real-world use of LLM generated materials (32). Previous studies consistently demonstrate that AI-generated patient education materials (PEMs) are written well above recommended public-health reading levels (Grade 6–8). For example, Beyoğlu, Kaya (33) reported that ChatGPT-generated cancer education materials required a Flesch–Kincaid Grade Level (FKGL) of 15.11 ± 7.36 and a Gunning Fog Index of 18.16 ± 7.40, implying that university-level reading skills are needed for comprehension .Similarly, Akkan and Seyyar (23)found that PEMs on fragility fractures generated by ChatGPT exhibited FKRE scores of 23.60–34.35 and FKGL values of 12.05–14.50, again exceeding the recommended range. When compared with these benchmarks, the outputs generated in our study demonstrate similar or even higher levels of complexity. GPT-5 produced highly technical responses, with a SMOG Index of 25.4 [95% CI: 23.7–27.0], significantly higher than the 14–18 range commonly reported in prior studies.

DeepSeek V3 showed the largest variability (FKRE = 5.2–27.6), shifting between clinical and conversational styles, while Grok-4 delivered the most consistent readability (FKRE = 23.6–32.2), although still above ideal levels for non-specialist users. This will greatly constrain the further application of these models. Developing responses that are accessible and easily understood may become a key focus of future research.

### Implications and Future Research

This study highlights both the potential and the limitations of LLMs in generating health-related content on ADHD. First, while all three models demonstrated alignment with DSM-5 in defining ADHD and describing its symptoms, their distinct emphases underscore how different design choices shape the scope and style of outputs. This suggests that model selection should be tailored to specific use cases: Grok-4 for clinically stable and structured responses, DeepSeek V3 for broad contextual guidance, and GPT-5 for concise and strengths-based perspectives. Second, the findings demonstrate that stability of response generation is varied between models. Grok-4’s relatively high stability positions it as a promising candidate for educational and clinical support contexts where consistency is vital, though its subscription model raises concerns about cost and accessibility. Conversely, DeepSeek V3 offers open access but with significant variability, which may limit reliability. Third, readability emerged as a persistent barrier across all models, with outputs exceeding recommended public-health literacy levels. This reinforces concerns raised in prior research that LLM-generated patient education materials may be inaccessible to the general public. Without improved accessibility, the broad dissemination of LLM-generated health information risks exacerbating health inequities.

Future research should investigate strategies to optimize the readability of LLM-generated health content, ensuring alignment with recommended public health literacy standards. Comparative studies across different languages and cultural contexts are needed to evaluate whether stability and complexity patterns are consistent beyond English-language outputs. Moreover, systematic experiments on fine-tuning, prompt engineering, and adaptive decoding strategies may help to balance stability with accessibility. Finally, longitudinal studies examining user comprehension, trust, and behavioral outcomes when interacting with LLM-generated materials would provide critical evidence for their safe integration into clinical and educational settings.

### Strengths and limitations

This study’s primary strength lies in its comparative design, which systematically tested three of the most advanced LLMs (ChatGPT-5, DeepSeek V3, Grok 4) using identical prompts and conditions. By incorporating multiple readability and lexical complexity indices, the analysis provided nuanced insights into both inter-model differences and intra-model variability. These findings offer practical guidance for different user groups teachers, parents, and healthcare providers who may have diverse needs when consulting AI-generated information about ADHD.

However, several limitations must be acknowledged. First, the study was conducted exclusively in English within a Melbourne-based testing environment, limiting generalizability to non-English-speaking populations. Second, the prompts were predefined and not iteratively adjusted, which may have constrained the models’ capacity to generate adaptive responses. Finally, the study focused primarily on textual readability and stability, rather than evaluating real-world comprehension or decision-making by specific user groups such as PE teachers.

## Conclusion

In summary, this study demonstrates that while all three LLMs aligned with DSM-5 in defining and describing ADHD, they differed markedly in scope, stability, and readability. Grok-4 showed the greatest stability, DeepSeek V3 offered the broadest contextual detail, and GPT-5 emphasized accessibility and strengths-based perspectives. However, the consistently high reading levels across models highlight a major barrier to practical application, underscoring the need for future work on improving accessibility and tailoring outputs to non-specialist audiences.

**Fig 1.**
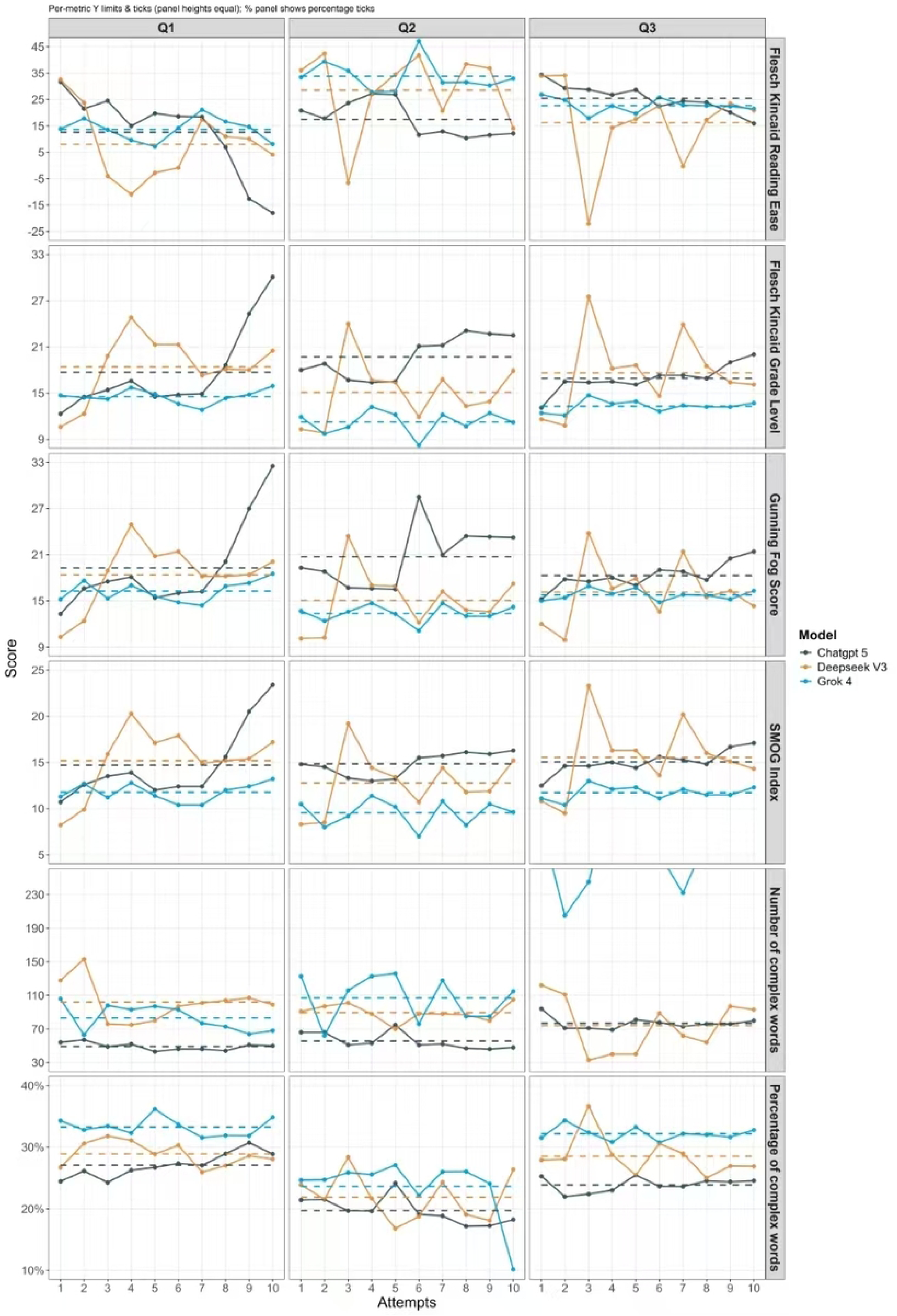
The chart shows readability trends of three language models (ChatGPT-5, DeepSeek V3, and Grok 4) across three questions.

## Data Availability

All relevant data are within the manuscript and its Supporting Information files.

## Declarations

### Ethics approval and consent to participate

Not applicable. This study did not involve human participants, and all data analysed were generated by the AI model.

### Availability of data and materials

The datasets generated and/or analysed during the current study are available from the corresponding author on reasonable request.

### Author contributions

**XMH:** Formal analysis, Writing – Review & Editing; **MZ:** Conceptualization, Methodology, Software, Formal analysis, Writing – Original Draft, Visualization; **RX:** Writing – Review & Editing.

## Acknowledgements

The authors gratefully acknowledge the developers of ChatGPT-5; DeepSeek V3; Grok-4 for providing the model used in this study, and thank all researchers whose prior work informed this evaluation.

## List of abbreviations

ADHD: Attention-deficit/hyperactivity disorder
AI: Artificial Intelligence
FKRE: Flesch–Kincaid Reading Ease
FKGL: Flesch–Kincaid Grade Level
LLM: Large Language Model
PEMs: Patient Education Materials
SMOG: Simple Measure of Gobbledygook
IEP: Individualised Education Program
PEMs: patient education materials

